# Evaluation of rotavirus, pneumococcal conjugate and human papillomavirus vaccination in four Pacific island countries: A cost-effectiveness modelling study

**DOI:** 10.1101/2025.04.14.25325775

**Authors:** Natalie Carvalho, Emma Watts, Victoria L. Oliver, Andrew Clark, Murat Ozturk, Siale Akauola, Clare Whelan, Take Naseri, Kylie Jenkins, Inez Mikkelsen-Lopez, Ki Fung Kelvin Lam, Rommel Rabanal, Ross McLeod, Mark Jit, Fiona M Russell

**Affiliations:** Melbourne School of Population and Global Health, University of Melbourne, 207 Bouverie St, Carlton, VIC 3053 Australia; Asia-Pacific Health, Murdoch Children’s Research Institute, Melbourne, Australia; London School of Hygiene and Tropical Medicine, Keppel Street, London, United Kingdom; UNICEF Pacific Islands Office; Ministry of Health, Tonga; Ministry of Health, Tuvalu; Ministry of Health, Samoa; Telethon Kids, Perth; Asian Development Bank; eSYS Development, Australia; Department of Paediatrics, The University of Melbourne

**Keywords:** Vaccination, Cost-effectiveness, Rotavirus, Pneumococcal, Cervical cancer, Pacific, Budget impact analysis, Fair pricing, New vaccine introduction

## Abstract

**Background:** To assist decision making on the introduction of rotavirus vaccine (RVV), pneumococcal conjugate vaccine (PCV) and human papillomavirus vaccine (HPVV), cost-effectiveness and budget impact evaluations were undertaken in Samoa, Tonga, Tuvalu and Vanuatu.

**Methods:** A proportionate outcomes model was used to evaluate vaccine introduction in each country from a health systems perspective, using country-specific data supplemented with regional and global estimates. A 10-year vaccination program was modelled from 2021, with costs and outcomes (disability-adjusted life years; DALYs) summed over a life-time horizon and discounted at 3%. Vaccine dose costs were based on Pan American Health Organization (PAHO) Revolving Fund prices, with lower priced products also explored.

**Findings:** Introduction of all three vaccines in all countries could prevent over 1,000 deaths over the lifetimes of the vaccinated cohorts. The cost per DALY averted at PAHO Revolving Fund prices ranged from 43% - 73% of the per capita gross domestic product (GDP) in each country, and 15% - 58% for lower-priced vaccines. The budget impact ranged from 359% (Samoa) to 1,368% (Vanuatu) of the 2019 vaccine budgets, and 149% (Samoa) to 775% (Vanuatu) for lower-priced vaccines. Cost-effectiveness results were most sensitive to disease burden, discount rate, vaccine efficacy, and program costs.

**Interpretation:** Development partner-supported introduction of HPVV, PCV and RVV may represent good value for money in Samoa, Tonga, Tuvalu and Vanuatu, depending on willingness to pay thresholds, but will place considerable burden on immunisation budgets. Financial sustainability requires increases in immunisation budgets and negotiation of affordable vaccine prices.

**Funding:** Asian Development Bank.

## Introduction

Despite substantial global progress, diarrhoea and lower respiratory tract infections remain leading causes of mortality in children aged 1 – 59 months, resulting in an estimated 1·22 million deaths in 2019.^1^ In the Pacific region (excluding Australia and New Zealand), lower respiratory tract infections are the leading cause of mortality in children under five (accounting for 26·4% of deaths), while diarrhoea is the fourth leading contributor to under five mortality (accounting for 9·9% of deaths).^2^ Cervical cancer is the fourth leading cause of cancer incidence and cancer-related mortality among women worldwide, causing an estimated 311,000 deaths in 2018, the majority occurring in resource-limited settings.^3,4^ In the Pacific region, rates of cervical cancer are some of the highest in the world and likely to be under-reported, due in part to inadequate coverage of screening programs.^5^ For example, cervical cancer is the leading cause of cancer death among women of reproductive age in Vanuatu, which has the lowest globally recorded average age at cervical cancer death of 45 years.^4,6^

Effective vaccines against these diseases have existed for 10-20 years and have formed part of comprehensive immunisation programs globally. The World Health Organization (WHO) recommends all infants be routinely immunised with rotavirus vaccine (RVV)^7^ to protect against rotavirus disease, which causes over a third of diarrhea-related mortality.^8^ Pneumococcal conjugate vaccines (PCV) are also recommended by the WHO to be administered to all children for the prevention of pneumococcal disease, a leading cause of lower respiratory tract infections in children.^9^ A vaccine against human papillomavirus (HPVV), the predominant cause of cervical cancer, is recommended for administration to girls aged nine years and over as part of WHO’s Cervical Cancer Global Elimination Strategy.^10^ All three vaccines are part of the immunization programs in many countries in the Asia Pacific region, including Australia, New Zealand, Fiji, Indonesia and the Philippines; however, at the time of this analysis were yet to be introduced in nine Pacific Island countries,^11^ despite WHO recommendations that this should be a priority for national immunization programs.^12^ In 2012, Fiji was the first country to successfully adopt all three vaccines together. Evidence from both Fiji and Kiribati shows a decline in morbidity and mortality due to severe diarrhoea, a decrease in childhood pneumonia hospital admissions in Fiji following vaccine introduction, and high effectiveness of HPVV against HPV detection in Fiji.^13–16^

Traditional childhood vaccines have been considered among the most cost-effective public health interventions.^17^ While newer vaccines tend to be more expensive, lower-priced vaccines have recently become available and received WHO pre-qualification, providing the opportunity for new vaccines to be cost-effective. With advances in technology and public health recommendations, countries are expanding their routine immunization schedules with resulting impacts on fiscal space for health. Organisations such as Gavi, the Vaccine Alliance, the Pan American Health Organization (PAHO) and UNICEF have supported low-income countries to introduce new vaccines through shared financing arrangements and pricing negotiations.

Until recently, Pacific Island countries have lagged in new vaccine introduction. Many Pacific Island countries are characterized by having small and highly dispersed populations, together with limited resources, remoteness, susceptibility to natural disasters and other external shocks, which increase dependence on external support for financing of immunisation programs (see Box 1 for further detail). Most Pacific middle-income countries are ineligible for Gavi support and, due to small population sizes, would lack the bargaining power to negotiate affordable vaccine prices on their own. However, the UNICEF Vaccine Independence Initiative (VII) has been serving to Pacific Island Countries with demand consolidation and bridge financing support.^18^ In 2018, a unique window of opportunity arose when development partner assistance through the Asian Development Bank (ADB) became available to fund vaccine introductions in Samoa, Tonga, Tuvalu and Vanuatu through a five-year project and cost-sharing arrangement, in the context of health systems strengthening.^19^ The specific vaccines selected for consideration of introduction by the four countries were HPVV, RVV and PCV, which were chosen based on successful introduction of these vaccines in Fiji, the strong evidence base for disease reduction, and the need to prevent cervical cancer. The project leveraged the VII, enabling access to vaccine prices similar to those obtained by PAHO Revolving Fund for Access to Vaccines,^20^ but much more expensive than Gavi prices. In this study, we estimate the cost-effectiveness and budget impact of simultaneous roll-out of HPVV, RVV and PCV in Samoa, Tonga, Tuvalu and Vanuatu. Results from this analysis formed part of the package of evidence informing the decision to adopt all three vaccines in each of the four countries. Implementation of these vaccines in each country was delayed due to the COVID-19 pandemic but began in 2021/2022.

## Methods

### Study design

We conducted a cost-effectiveness and budget impact analysis of introducing three new vaccines in four Pacific Island countries: Samoa, Tonga, Tuvalu and Vanuatu. Vaccines included: RVV, delivered to children under 5 by a two-dose schedule (RV1) schedule alongside diphtheria-tetanus-pertussis (DTP)-containing vaccine doses 1 and 2; PCV (PCV13), administered to children under 5 by a three-dose schedule alongside DTP-containing vaccine doses 1-3; and one dose of HPVV (bivalent HPV2or quadrivalent HPV4), administered to ten-year-old girls alongside existing school-based vaccine programs. In a separate analysis, we also evaluated lower price vaccines for RVV (a three-dose schedule), PCV (three dose schedule of PCV10) and HPVV (one dose schedule of bivalent HPV2) that have more recently become available on the market. There is insufficient evidence to support modelling of differential efficacy across the different brands of vaccines and where possible this analysis was designed to be brand-agnostic in line with WHO position papers and UNICEF policies supporting transparent and competitive procurement principles. We used input parameters (such as vaccine cost and efficacy) that were applicable to both PCV10 and PCV13; as well as HPV2 and HPV4 vaccine.

A ten-year vaccination program (2021-2030) was modelled assuming simultaneous introduction of all three vaccines into routine immunisation programs (without a catch-up campaign), compared to no vaccine. We used UNIVAC (version 1·4), an Excel-based proportionate outcomes model that has been used extensively around the world to evaluate the cost-effectiveness of vaccines in low- and middle-income countries (LMICs).^21–25^ The model has been specifically designed for ease of use at country level,^26^ and it provides a basis for strengthening national capacity where feasible, building consensus between stakeholders and increasing the local ownership and policy-relevance of results. See Webappendix A for an overview of model structure and how inputs are combined to produce outputs. Each vaccine was modelled separately in UNIVAC (compared to the status quo) and results were combined to provide estimates of the joint vaccine introduction scenario.

We used a healthcare payer perspective in the base case, including both government and development partner costs, and considered a partial societal perspective that included household out-of-pocket costs in a separate base-case scenario. All costs were adjusted to 2019 United States dollars (USD). Costs in other currencies were first exchanged to local currency units (LCU) using World Bank year-specific exchange rates. Costs in LCU were inflated to 2019 LCU using country-specific World Bank gross domestic product (GDP) deflators, then exchanged to 2019 USD using World Bank USD/LCU 2019 exchange rates.^27^

Where available, country-specific data on disease burden were used as model inputs. However, due to a scarcity of local data, data were predominantly sourced from published studies undertaken in other Pacific Island countries (particularly Fiji) or informed by global estimates. Rotavirus and pneumococcal disease events (cases, outpatient visits, hospitalisations and deaths) and treatment costs were estimated for successive birth cohorts over the first five years of life while cervical cancer events (cases, hospitalizations, and deaths) and treatment costs were estimated over the entire lifetime of a woman, with and without each vaccine. Average life expectancy, duration of illness and disability weights were used to estimate disability-adjusted life years (DALYs) over a life-time perspective, with future costs and benefits discounted at 3% in the base case.

### Disease burden

Inputs used for modelling disease burden are presented in Table 1 and in the Supplementary material (Webappendix B).

**Table 1:**
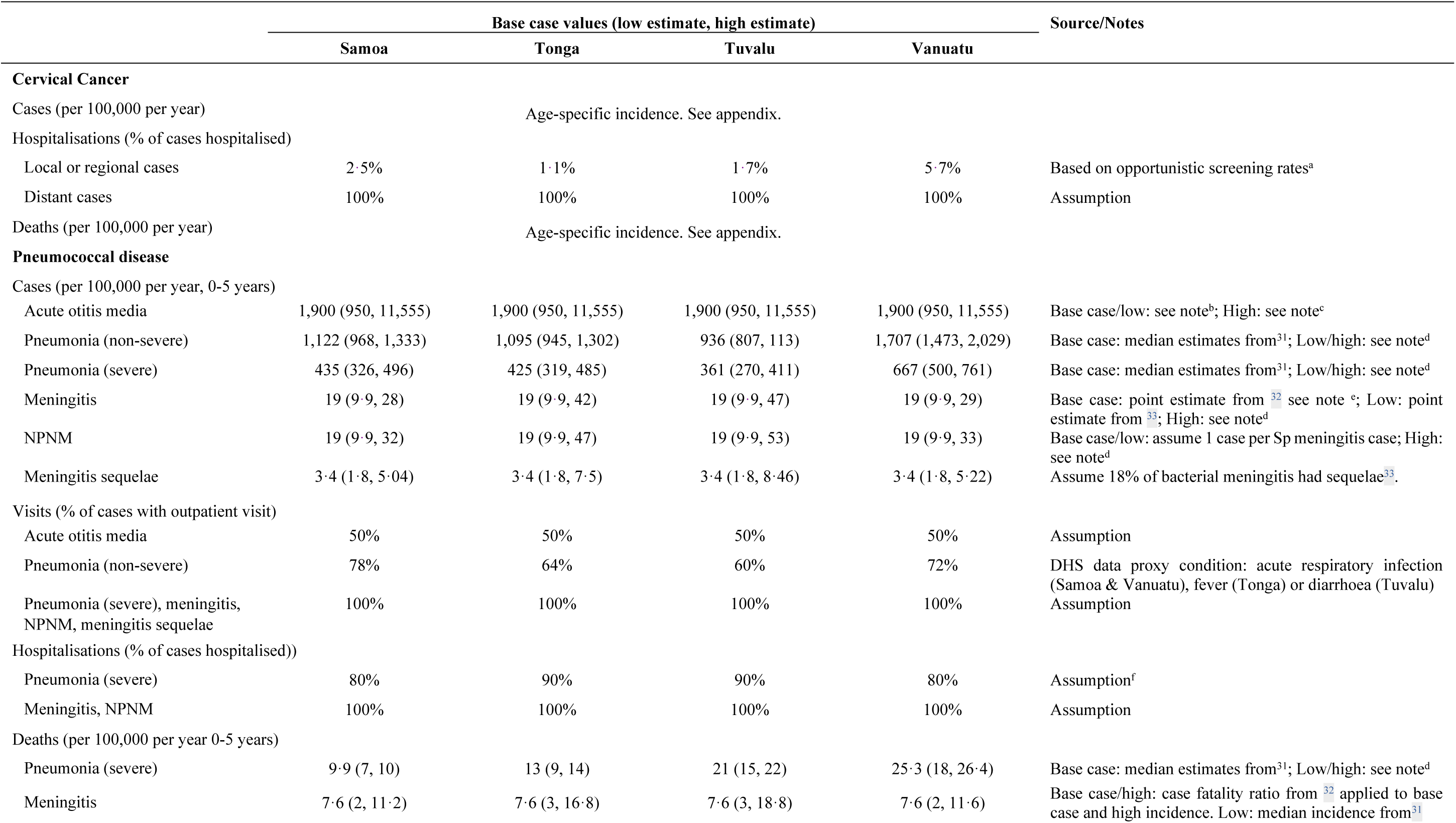
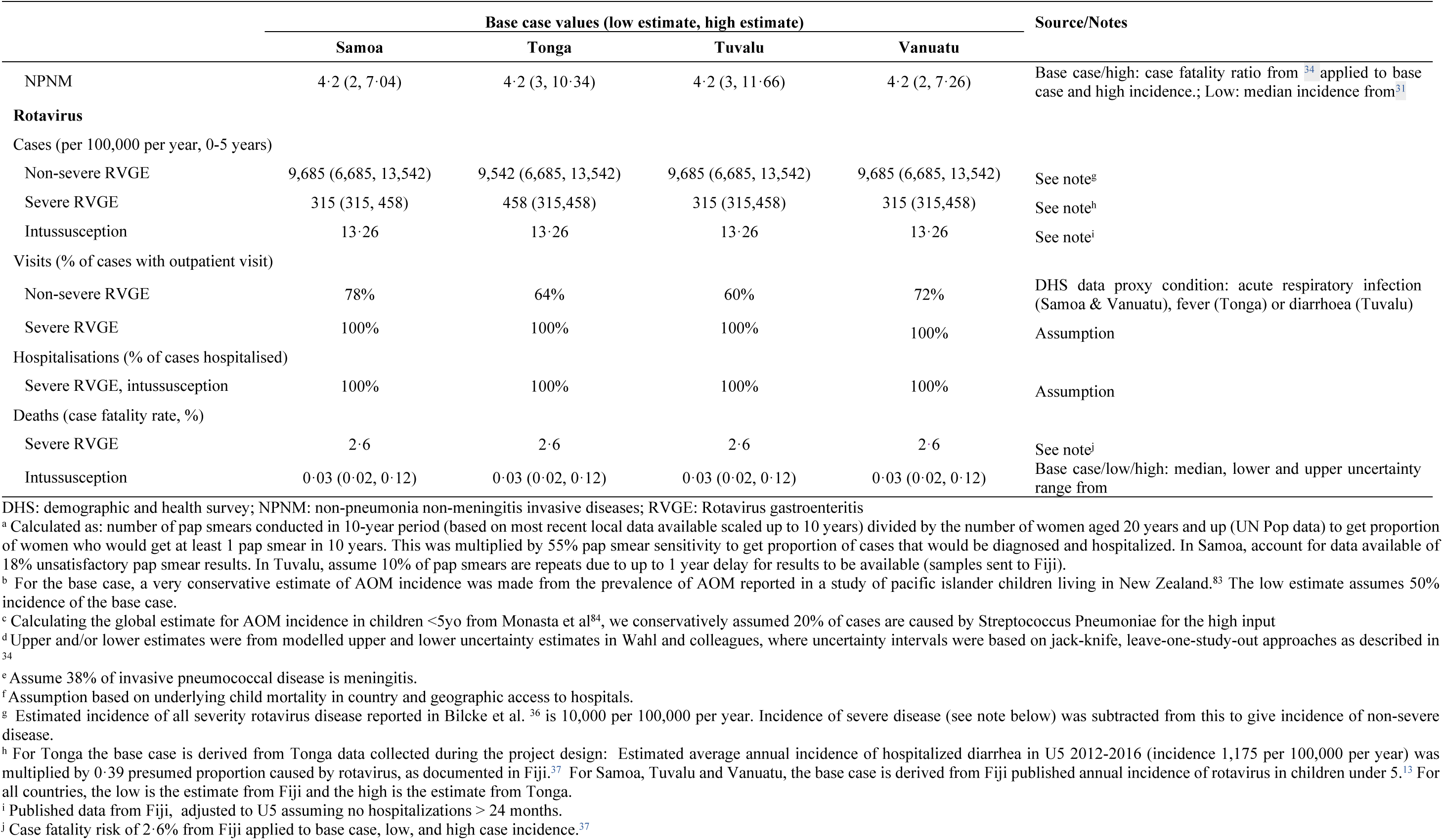
Inputs for modelling disease burden. Table gives base case for inputs used to model cases, visits, hospitalisation and deaths. Where applicable, inputs for low and high estimates in parentheses.

### Cervical cancer disease incidence and mortality

Due to limited country-specific data at the time of this analysis, age-specific incidence and mortality rates of cervical cancer in all countries were assumed to be the same as a published study on age-specific incidence of cervical cancer in Melanesian Fijian women using actual cases rather than estimates from models (Webappendix B, Table B2).^6^ These rates were mostly higher than more recent GLOBOCAN estimates that have become available for Samoa and Vanuatu,^4^ and higher than the most recent Global Burden of Disease (GBD) estimates.^28^ We assumed a given distribution of incident cases across severity levels (19% local, 73% regional, and 9% distant incident cases), based on a recent global analysis that accounted for information about staging of cervical cancer cases in LMICs.^29^

### Pneumococcal disease incidence and mortality

The model includes six health states and outcomes associated with pneumococcal disease including: acute otitis media, non-severe and severe pneumonia, meningitis, other non-pneumonia non-meningitis invasive diseases (NPNM) and meningitis sequelae.

In the base case, the incidence of acute otitis media is based on data on Pacific Island children living in New Zealand and assumed to be the same across all countries.^30^ For the incidence of severe and non-severe pneumonia, we used country-specific estimates derived from the WHO and Maternal and Child Epidemiology Estimation collaboration.^31^

For the incidence of meningitis, we used data from a study of invasive pneumococcal disease in children under the age of five in Tonga between 2010 and 2013, where 38% of cases were meningitis.^32^ This was applied to all countries. We assumed one case of non-pneumonia non-meningitis invasive disease per case of meningitis for the base case. We assumed 18% of bacterial meningitis cases were associated with sequelae based on a study of clinical manifestations of invasive pneumococcal disease in children under five in Fiji.^33^

For the incidence of pneumococcal pneumonia deaths, we used country-specific estimates from WHO and Maternal and Child Epidemiology Estimation collaboration.^31^ A case fatality ratio of 40% for meningitis was used in the base case from a published study of invasive pneumococcal disease in Tonga.^32^ A case-fatality ratio of 22% was used for severe non-pneumonia non-meningitis invasive diseases based on a published estimate for the Western Pacific region.^34^ The age distribution of pneumococcal disease events in each week of age under five years was based on a global review (Webappendix B, Table B3).^35^

### Rotavirus disease incidence and mortality

Cases of non-severe rotavirus gastroenteritis in children under five years old in each country were based on a global systematic review of symptomatic rotavirus infections in children.^36^ For Tonga, the estimate for hospitalised rotavirus gastroenteritis was based on local data estimating the incidence of hospitalised gastroenteritis in children under five, assuming 39% of gastroenteritis hospital admissions were attributed to rotavirus as was found in Fiji.^13^ For Samoa, Tuvalu and Vanuatu, the estimate was based on the Fiji study of incidence of hospitalised rotavirus gastroenteritis in children under five, in the years prior to vaccine introduction as no local data were available.^13^ We estimated that the incidence of severe rotavirus gastroenteritis was equal to the incidence of hospitalised cases.

Evidence of the case fatality ratio of 2·6% for hospitalised rotavirus gastroenteritis in children under five in Fiji was applied to the incidence of cases to estimate mortality in all countries.^37^ The age distribution of rotavirus disease events in each week of age under five years was estimated from a global review and statistical analysis of 92 hospital admission datasets, stratified by child mortality strata.^38^

### Intussusception

Rarely, RVV causes a serious adverse event, intussusception, which requires hospitalisation and treatment. The baseline incidence of intussusception was applied from a published study in Fiji, prior to RVV introduction,^39^ adjusting the reported rate in children under two years of age in Fiji to annual incidence of intussusception per 100,000 children under five in each country (Webappendix B, Table B3). We assumed a case fatality ratio of 0·03%, based on a meta-analysis estimating the case fatality ratio for the Western Pacific region.^40^ We estimated a relative risk of intussusception associated with RVV in the 1-7 day risk period based on a recent meta-analysis.^41^ The age distribution of intussusception cases and deaths was based on a recent global review (Webappendix B, Table B3).^40^

### Disability weights, duration and calculations of disability-adjusted life years

To estimate years lived with disability, country-specific demographic data and life expectancy at birth were sourced from the United Nations Populations Projections (Webappendix B, Table B1). For non-fatal conditions, years lived with disability were based on disability weights and duration of illness. Disability weights for all vaccine-preventable diseases across all countries came from the GBD Study 2013 (Webappendix B, Table B4).^42^

The average duration of illness for local, regional and distant cervical cancer was based on the 2017 GBD study estimates.^43^ For pneumococcal disease, we assumed an average duration of seven days for acute otitis media and non-severe pneumonia, and a duration of 10 days for severe pneumonia, meningitis and other severe, non-pneumonia non-meningitis disease. We assumed an average duration of 50 years for meningitis sequelae. For rotavirus, we used an average duration of three days for non-severe and seven days for severe gastroenteritis,^44^ and assumed a duration of seven days for intussusception.

### Vaccine coverage, timeliness and efficacy

Inputs used to model vaccine coverage and efficacy are presented in Table 2 and Table 3. In the status quo we model no coverage of PCV, RVV and HPVV. For the intervention, HPVV coverage assumptions were based on 2023 WHO/UNICEF Estimates of National Immunization Coverage (WUENIC) data on HPVV coverage in Fiji, where HPVV has been a part of the routine immunization schedule for several years.^45^ The model was updated using country-specific data on actual HPVV coverage in 2023 (following vaccine introduction) in the lower estimate for the sensitivity analysis. Routine coverage of RVV and PCV were based on country-specific coverage from WUENIC 2023 using DTP-containing vaccine doses 1-3 as proxy, given that these vaccines are given to infants together in a bundle.

**Table 2:** Vaccine coverage estimates.

|  | Base case values (low estimate, high estimate) |  |  |  |
| --- | --- | --- | --- | --- |
|  | Samoa | Tonga | Tuvalu | Vanuatu |
| HPVV<sup>a</sup> |  |  |  |  |
| Dose 1 | 87·0 (86·0, 87·0) | 86·0 (30·0, 86·0) | 86·0 (84·0, 86·0) | 86·0 (41·0, 86·0) |
| PCV/RVV<sup>b</sup> |  |  |  |  |
| Dose 1 | 100·0 (90·0, 100·0) | 99·4 (89·5, 99·4) | 100·0 (90, 100·0) | 96·0 (86·4, 100·0) |
| Dose 2 | 91·9 (82·7, 91·9) | 99·3 (89·4, 99·3) | 97·0 (87·4, 97·0) | 93·0 (83·7, 95·2) |
| Dose 3 | 83·2 (74·8, 83·2) | 99·2 (89·3, 99·2) | 94·1 (84·7, 94·1) | 90 (81·0, 90·5) |

HPVV: Human papillomavirus vaccine; PCV: Pneumococcal conjugate vaccine; RVV: Rotavirus vaccine
<sup>a</sup> Except for Samoa, WUENIC data on coverage of first dose of HPVV among the target population in 2023 in Fiji was used for the base case and high estimate. The low estimate used country-specific WUENIC data on coverage of first dose of HPVV among the target population in 2023. In Samoa country-specific data was used in the base case and high, while coverage data from Fiji was used as the low estimate.
<sup>b</sup> Samoa, Tonga, and Tuvalu: Base case used official coverage estimates of DTP doses 1 and 3 (2023) from WHO/UNICEF and high used administrative coverage estimates (which in some cases were equivalent to official coverage estimates). For Vanuatu: due to a decline in immunization coverage following the COVID-19 outbreak, data from 2019 were used. For all countries, low estimate was 90% of base case and dose 2 coverage was taken as a linear average of dose 1 and 3 coverage due lack of data to inform plausible estimates for these parameters.

**Table 3:** Vaccine efficacy and duration estimates. Vaccine efficacy from 2 weeks after vaccination unless otherwise stated. The same efficacy and duration estimates were used for all countries unless otherwise stated.

|  | Base case values (low estimate, high estimate) | Source/Notes |
| --- | --- | --- |
| **HPVV** |  |  |
| Dose 1 efficacy (%) | 73·8 (59·0, 79·0) | See note<sup>a</sup>. |
| Duration of efficacy (years, all doses) | 100 | Assumption based on <sup>49,50</sup> |
| **PCV** |  |  |
| Dose 1 efficacy (severe disease, %)<sup>b</sup> | 29 (15, 38) | Assume dose 1 efficacy is 50% of dose 2 |
| Dose 2/3 efficacy (severe disease, %)<sup>b</sup> |  |  |
| 3 years after vaccination | 58 (29, 75) | Base case/low/high: median, lower and upper uncertainty range from <sup>52</sup> |
| 4 years after vaccination | 57 (28·5, 73·7) |  |
| 5 years after vaccination | 28·2 (14·1, 36·5) |  |
| Duration of efficacy (years, all doses) | 5 | Assumption based on <sup>54</sup> |
| **RVV (Samoa, Tonga, Tuvalu)** |  |  |
| Dose 1 efficacy (severe disease, %)<sup>c</sup> | 57·8 (45·5, 73·6) |  |
| Dose 2/3 efficacy (severe disease, %)<sup>c</sup> |  |  |
| 2 weeks after vaccination | 91·4 (89·8, 92·7) | Base case/low/high: median, lower and upper uncertainty range from <sup>55</sup> |
| 6 months after vaccination | 76·8 (72·7, 80·2) |  |
| 12 months after vaccination | 69·5 (64·3, 73·9) |  |
| **RVV (Vanuatu)** |  |  |
| Dose 1 efficacy (severe disease, %)<sup>c</sup> | 49·9 (38·2, 65·3) | Base case/low/high: median, lower and upper uncertainty range from <sup>55</sup> |
| Dose 2/3 efficacy (severe disease, %)<sup>c</sup> |  | Base/Low: median and lower uncertainty range from <sup>55</sup> High: see note<sup>e</sup> |
| 2 weeks after vaccination | 78·9 (75·5, 91·4) |  |
| 6 months after vaccination | 45·7 (38·2, 76·8) |  |
| 12 months after vaccination | 31·8 (23·8, 69·5) |  |

HPVV: Human papillomavirus vaccine; PCV: Pneumococcal conjugate vaccine; RVV: Rotavirus vaccine
<sup>a</sup> Base case: Assume 97·5% efficacy against high-risk subtypes based on median efficacy of one dose in a study in Kenya <sup>47</sup> multiplied by percent of cancer caused by high-risk types, taken to be 75·7% based on median estimate for Oceania <sup>85</sup>. Low: Assume 81·7% efficacy against high-risk subtypes (HPV16/18) based on lower bound 95% confidence interval in Kenya study<sup>47</sup> multiplied by lower bound estimate for percent of cancer caused by high-risk types reported for Oceania (72·3%) <sup>85</sup>. High: Assume 100% efficacy against high-risk subtypes based on , multiplied by upper bound 95% confidence interval for percent of cancer caused by high-risk types reported for Oceania (79·0%) <sup>85</sup>. multiplied by lower bound estimate for percent of cancer caused by high-risk types reported for Oceania (72·3%) <sup>85</sup>. High: Assume 100% efficacy against high-risk subtypes based on <sup>86</sup>, multiplied by upper bound estimate 95% confidence interval for percent of cancer caused by high-risk types reported for Oceania (79·0%) <sup>85</sup>.
<sup>b</sup> Efficacy against non-severe Sp pneumonia as a proportion of efficacy against severe disease = 0·07.<sup>53</sup>
<sup>c</sup> Efficacy against non-severe RVGE as a proportion of efficacy against severe RVGE = 0·85.<sup>56</sup>
<sup>e</sup> Base case efficacy estimates for Samoa, Tonga and Tuvalu used as high estimate for Vanuatu dose 2/3 efficacy.

The UNIVAC model can capture vaccine timeliness through age-specific coverage rates. Following discussions with in-country vaccine delivery personnel, routine childhood vaccines were assumed to be delivered on time for Samoa and Tuvalu, and approximately 90% of final dose-specific coverage rates achieved at the target age and final coverage achieved by two years of age for Tonga and Vanuatu. As a result, all RVV doses were assumed to be administered within the manufacturers’ recommended age window (i.e. the first dose within 15 weeks and the last dose within 32 weeks).

Vaccine efficacy estimates are from a detailed review of the existing literature - drawing on the most recent systematic reviews and randomised controlled trials as a priority – along with feedback from experts in the field. Estimates used for vaccine efficacy (from two weeks after vaccination) and the duration of protection were the same across all countries, except for RVV as Rotavirus vaccines perform better in settings with low rotavirus related mortality.^46^ For HPVV, we assumed a 73·8% vaccine efficacy. This was calculated by multiplying the efficacy of one dose against high-risk subtypes found in a large-scale, randomised controlled trial in Kenya (97·5%)^47^ by the estimated percent of cancer caused by HPV 16 and 18 in Oceania (75·7%).^48^ We assumed a lifetime duration of protection based on two previous systematic reviews of HPVV cost-effectiveness analyses.^49,50^ We assume this same efficacy for one dose of the low cost HPVV based on a recent interim analysis of a phase 3 clinical study.^51^ PCV efficacy against severe pneumonia with *Streptococcus pneumoniae* (Sp) as a causative pathogen was estimated at 58% for three doses based on a Cochrane systematic review.^52^ A conservative assumption of 4% PCV efficacy was estimated against non-severe Sp pneumonia following three doses.^53^ We assume a five-year duration of protection for PCV (the highest risk period for children) with waning starting in the last year.^54^ Initial vaccine efficacy for a full course of RVV against severe disease was estimated based on a recent meta-analysis of randomised controlled trials by child mortality strata and was estimated at 91·4% for Samoa, Tonga and Tuvalu, and 78·9% for Vanuatu (where child mortality is higher than the other three countries).^55^ The assumed waning of RVV efficacy was ￼ on the same analysis.^55^ Efficacy against non-severe rotavirus gastroenteritis was assumed to be 85% of the vaccine efficacy against severe disease.^56^ For PCV and RVV, efficacy and waning were assumed to be the same between the lower-price and higher-price products evaluated.

### Vaccine program costs

All vaccine dose and program costs are summarised in Table 4 and Appendix Table C4. Vaccine dose costs were informed by UNICEF based on PAHO Revolving Fund pricing (2017 price list), where we assumed a per dose price of $7·50 for RVV, $16 for PCV and $12 for HPVV in the base case analysis. These vaccine dose prices reflected the assumption that all four countries introduce all three vaccines to benefit from negotiated reduced prices. In a separate analysis, where we evaluated lower price vaccines, the per dose costs were assumed to be $1·15 for RVV and $4·00 for PCV based on UNICEF 2022 pricing, and $9·23 for HPVV based on World Health Organization Market Information for Access to Vaccines purchase database.

**Table 4:**
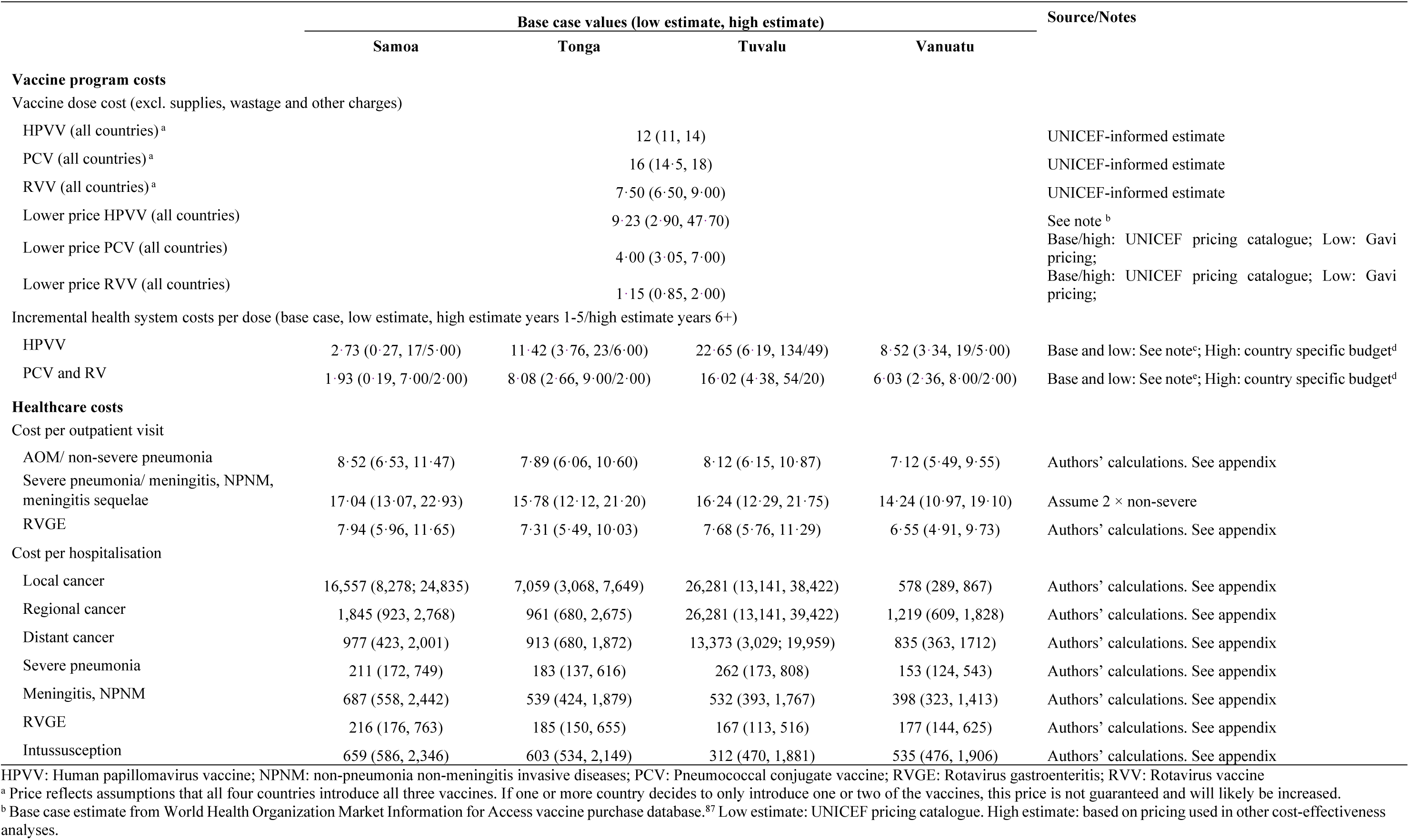
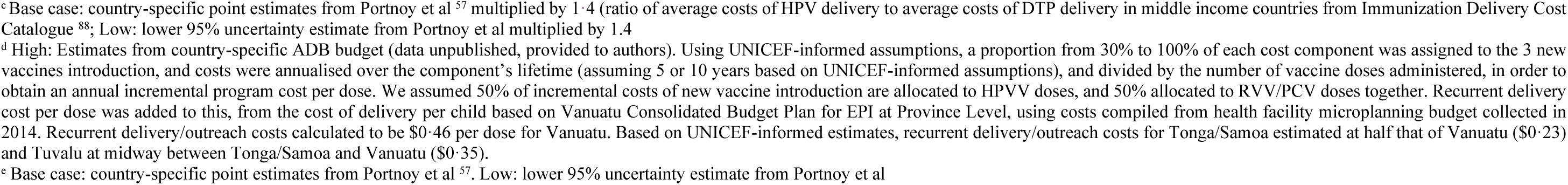
Program costs and healthcare costs (all in 2019 USD). See Appendix for supplies, wastage, and other costs applied to the vaccine dose costs.

The costs of other vaccine supplies (including syringes and safety boxes/bags), international handling and delivery charges, and wastage were based on UNICEF-informed estimates as outlined in Webappendix C (Table C1). We assumed a 10% wastage rate for single dose vial vaccines, whereas lower price RVV were assumed to be a 5-dose vial presentation with 80% wastage in Tuvalu due to its small population and 50% wastage in other countries based on expert opinion (wastage applied only to vaccine dose and not other consumables).

Incremental health systems costs per dose were included in the model to represent the additional cost to the health system of introducing new vaccines plus recurrent delivery costs (Table 4). For RVV and PCV, we used country-specific estimates from the published literature.^57^ These estimates were largely higher than those published in the Immunization Delivery Cost Catalogue,^58^ however are considered to more accurately reflect the costs by accounting for local contextual factors (such as GDP, population, and the existing routine vaccination schedule). For HPVV delivery costs we inflated the RVV and PCV delivery costs by a factor of 1·4 based on the difference in delivery costs between DTP3 and HPVV for middle income countries reported in the Immunization Delivery Cost Catalogue. We did not include any savings in incremental health system costs that might be possible from introducing multiple vaccines at once.

### Healthcare costs

Healthcare costs were from mixed sources, including national or regional-level data and costing studies, and the published literature. We used WHO-CHOICE (WHO Choosing Interventions that are Cost Effective) 2011 country-specific estimates for unit costs of hospital and outpatient visits and hospital bed days. Laboratory and pharmacy costs were from in-country fee schedules where available. Unit costs per visit and hospitalisation were moderated down by assumed rates of care-seeking (see Table 1 for care-seeking assumptions). Full details are available in Table 4 and the Supplementary material (Webappendix C).

A societal perspective was explored by including both direct costs (transportation and user-fees where applicable) and indirect costs (productivity losses for caregivers and women undergoing treatment for cervical cancer). Productivity losses were estimated by multiplying average length of hospital stay by country-specific female labour force participation rate from World Bank Data.^59^ Lost days of work were valued based on country-specific GDP per capita. Productivity losses due to death were not included in the model, and as a result our findings will represent a conservative estimation of the societal costs.

### Cost-effectiveness

A cost-effectiveness ratio, presented in terms of USD per DALY averted, was calculated for each vaccine compared to the status-quo (no vaccine), and a joint program cost-effectiveness ratio was calculated for introduction of all three vaccines together compared to the no vaccine scenario.

Because explicit country-specific or regional cost-effectiveness thresholds are not available in the Pacific, we compared cost-effectiveness ratios with two potential estimates of willingness to pay. As an upper estimate, we used willingness to pay thresholds of 1× the 2019 GDP per capita (Samoa $4,030; Tonga $3,749; Tuvalu $3,084; Vanuatu $2,861), based on historical recommendations of the WHO Commission on Macroeconomics and Health.^60,61^ As a lower estimate, we used recent willingness to pay estimates of 0·61×, 0·44× and 0·21× GDP per capital in Samoa, Tonga and Vanuatu, respectively.^62^ As estimates for Tuvalu were not available in this study, we used willingness to pay thresholds from Samoa as a proxy.

### Uncertainty analysis

We conducted one-way and probabilistic sensitivity analyses to account for the considerable uncertainty surrounding underlying epidemiological and cost data and test the robustness of our results to changes in key input parameters and model choices. Upper- and lower-bound estimates were explored across estimates of disease event rates - which included incidence and mortality - (Table 1), disability weights and durations (Webappendix B, Table B4; explored in the probabilistic sensitivity analysis only), vaccine coverage rates (Table 2), vaccine efficacy and duration (Table 3), and program and healthcare costs (Table 4 and Webappendix C).

For the one-way sensitivity analysis of discount rate, we ran a scenario with costs discounted at 3% and cost-effectiveness ratios calculated using undiscounted DALYs averted in line with recent WHO guidance on discount rates.^63^ We ran another scenario where both costs and DALYs were discounted at 6% (ADB-recommended discount rate for social sector projects).

For the probabilistic sensitivity analysis, there were limited data to inform the true underlying distributions for most parameters and the extent to which each of the parameters are correlated. We therefore assumed simple PERT-Beta distributions for each parameter as others have done with UNIVAC,^26,64^ and ran the model 1,000 times for individual vaccines in each country. Although this is a simplification, the PERT-Beta distribution is able to reproduce plausible distributions that reflect the mid/low/high values specified for each parameter, including right-skewed and left-skewed distributions. Dose prices were fixed in probabilistic analyses.^65^ For each draw, costs were summed across the three vaccines and likewise for DALYs averted to give results for the full program. Results from each draw were directly plotted on cost-effectiveness planes to generate cost effectiveness clouds rather than point estimates (Figure 1). To generate cost-effectiveness acceptability curves representing the probability of the program being cost effective at different willingness to pay thresholds, the proportion of draws with cost-effectiveness ratios below increasing willingness to pay thresholds, with thresholds starting from 10 USD per DALY averted and increasing in 10 USD increments until probability approached 1.

**Figure 1:**
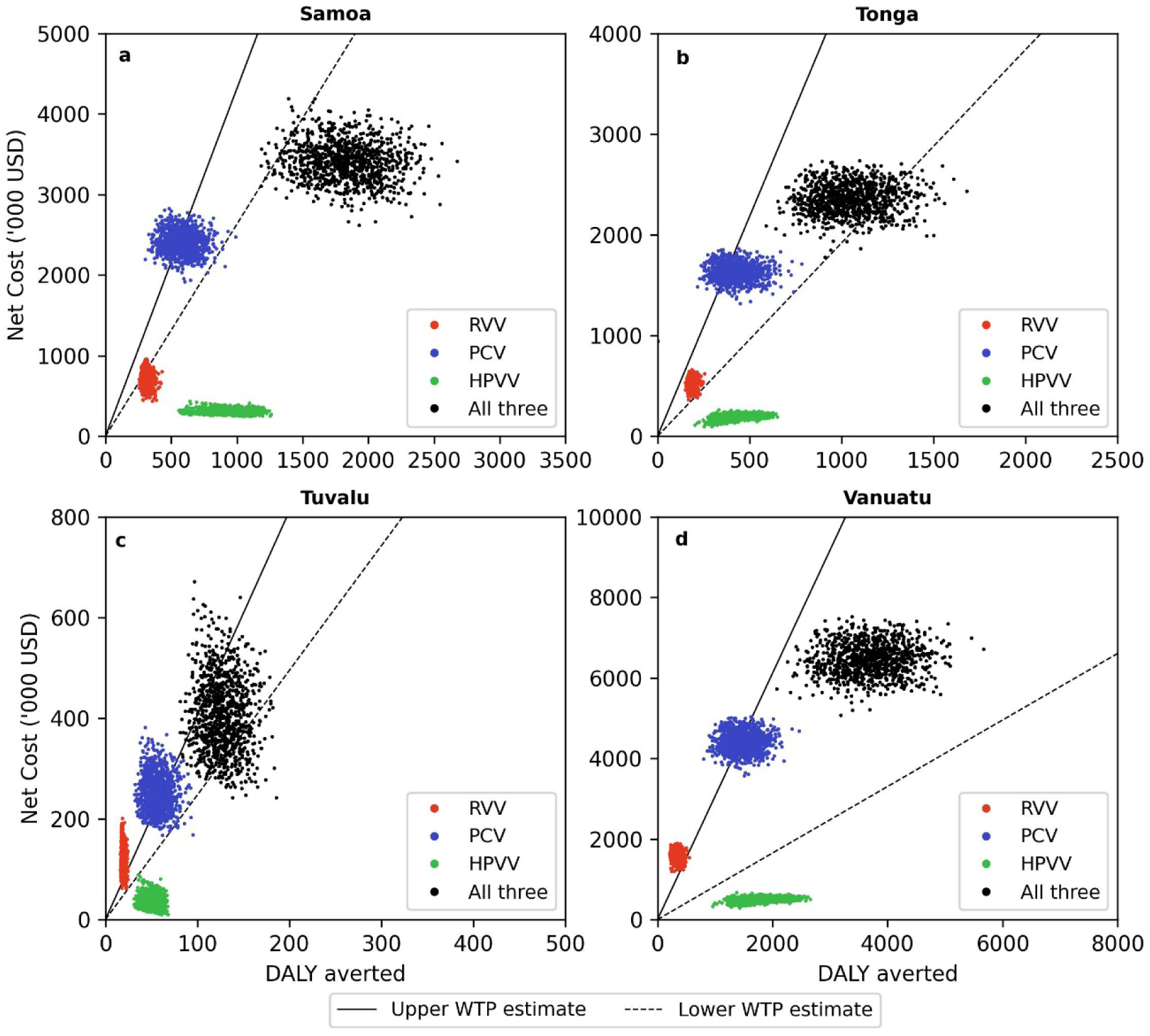
Cost per DALY averted results from probabilistic sensitivity analysis of HPVV, PCV and RVV in a: Samoa; a: Tonga; c: Tuvalu; and d: Vanuatu. Solid black line: Upper WTP estimate is 1× GDP per capita in each country. Dashed black line: Lower WTP estimate is 0·61×, 0·44×, 0·61×, and 0·21× GDP per capital in Samoa, Tonga, Tuvalu and Vanuatu, respectively. DALY: Disability-adjusted life year; HPVV: Human papillomavirus vaccine; PCV: Pneumococcal conjugate vaccine; RVV: Rotavirus vaccine; USD: United States Dollars; WTP: Willingness to pay

### Budget impact analysis

We estimated the budget impact of the new vaccine introductions from a government perspective, factoring in development partner support. Government share of dose costs began at 0% in year one and increased by 20 percentage points each year over the first five years until the full vaccine dose cost is paid by the government. Over the first five years, we modelled 50% of the incremental health system costs of vaccine introduction to be paid by government, increasing to 100% for years six to ten. We present annual undiscounted vaccine program costs for the base case and lower price vaccines separately by country and vaccine, alongside averted healthcare costs over a ten-year period. Both financial (program costs) and economic (healthcare costs averted) costs were included in the budget impact analysis. We also present results in terms of increase in total health expenditure and immunisation budget (undiscounted) based on budget impact at year six when the full cost of the program is paid by the government.

## Results

### Base case scenario

Vaccine program costs, healthcare cost savings to government, and health benefits of a 10-year vaccine program adopting HPVV, PCV and RVV at PAHO Revolving Fund pricing are shown in Table 5. The costs per DALY averted (compared to the status quo) for these vaccines are shown in Figure 1. See Webappendix D (Table D1 and Figure D1) for the same data for the lower price vaccines.

**Table 5:** Base-case cost-effectiveness results for HPVV, PCV and RVV. Cost-effectiveness ratios for each vaccine and all three vaccines are expressed as cost per DALY averted compared to the status quo (10-year program, lifetime benefit stream).

|  | Samoa | Tonga | Tuvalu | Vanuatu |
| --- | --- | --- | --- | --- |
| **HPV** |  |  |  |  |
| Vaccine program costs (USD) | 313,380 | 249,570 | 36,385 | 618,217 |
| Healthcare cost savings (USD) | 20,336 | 6,171 | 9,849 | 31,803 |
| Hospitalisations averted | 43 | 22 | 2 | 129 |
| Deaths averted | 226 | 124 | 13 | 530 |
| DALYs averted | 1,003 | 556 | 58 | 2,267 |
| CER (USD/DALY averted) | 292 | 437 | 456 | 257 |
| **PCV** |  |  |  |  |
| Vaccine program costs (USD) | 2,611,339 | 2,020,523 | 259,463 | 5,239,775 |
| Healthcare cost savings (USD) | 124,893 | 66,627 | 7,764 | 204,474 |
| Outpatient visits averted | 1,878 | 1,987 | 106 | 3,672 |
| Hospitalisations averted | 488 | 312 | 28 | 1,204 |
| Deaths averted | 23 | 16 | 2 | 60 |
| DALYs averted | 622 | 421 | 56 | 1,582 |
| CER (USD/DALY averted) | 3,997 | 4,646 | 4,479 | 3,184 |
| **RVV** |  |  |  |  |
| Vaccine program costs (USD) | 922,469 | 822,047 | 121,689 | 2,061,196 |
| Healthcare cost savings (USD) | 167,579 | 100,768 | 7,667 | 128,773 |
| Outpatient visits averted | 10,736 | 5,227 | 501 | 9,560 |
| Hospitalisations averted | 502 | 424 | 30 | 482 |
| Deaths averted | 11 | 6 | 1 | 10 |
| DALYs averted | 297 | 176 | 18 | 265 |
| CER (USD/DALY averted) | 2,542 | 4,102 | 6,300 | 7,284 |
| **All three vaccines** |  |  |  |  |
| CER (USD/DALY averted) | 1,839 | 2,531 | 2,972 | 1,836 |
| CER as a percentage of GDP (2019) | 43% | 58% | 73% | 60% |
| Societal CER (USD/DALY averted) | 1,808 | 2,500 | 2,950 | 1,801 |
| Societal CER as a percentage of GDP (2019) | 42% | 57% | 73% | 59% |

CER: Cost-effectiveness ratio; DALY: Disability-adjusted life year; GDP: Gross domestic product; HPV: Human papillomavirus vaccine; PCV: Pneumococcal conjugate vaccine; RVV: Rotavirus vaccine; USD: United States Dollars

Simultaneous introduction of all three vaccines in all four countries could prevent 1,022 deaths over the lifetimes of the people vaccinated in the ten-year program (893 cervical cancer deaths, 101 pneumococcal deaths, 28 rotavirus deaths), equivalent to just over half of the deaths that would otherwise have been caused by these diseases. Over 33,000 outpatient visits and 3,600 hospitalisations would be averted across the four countries. Due to the assumption of timely administration of vaccines in each country, excess cases of intussusception would be negligible (<1 over the ten- year vaccination program).

In the base case analysis, the cost-effectiveness ratio for HPVV falls just at or below 0·10× of GDP per capita in all countries. The cost-effectiveness ratios for PCV ad RVV mostly fall between 1× and 1·5× GDP per capita in all countries except for RVV in Samoa (cost-effectiveness ratio of 0·59× GDP per capita) and RVV in Vanuatu (cost-effectiveness ratio of 2·3× GDP per capita).

Considering all three vaccines introduced together at PAHO Revolving Fund prices, the joint cost-effectiveness ratio (compared to the current status with no vaccines) ranged from 0·43× GDP per capita in Samoa to 0·73× GDP per capita in Tuvalu. The secondary analysis conducted from a societal perspective resulted in slightly more favourable results (lower cost-effectiveness ratios) but minimal difference in findings.

Cost-effectiveness ratios of the lower price vaccines were mostly more favourable than for the PAHO Revolving Fund -priced vaccines, ranging from 0·05× GDP per capita (for HPVV in Samoa) to 1·9× GDP per capita (for RVV in Tuvalu). The cost-effectiveness ratio for the lower-priced RVV was higher than for the higher-price product owing to the three-dose schedule and the high per dose delivery costs in Tuvalu. The simultaneous introduction of lower-priced vaccines had a cost-effectiveness ratio ranging from 0·15× GDP per capita in Samoa to 0·58× GDP per capita in Tuvalu.

### Sensitivity analysis

Tornado diagrams for the one-way sensitivity analysis are shown in Figure 2 and in Webappendix D (Figure D2) for the lower price vaccines. Due to the preventative nature of these vaccine programs, with health benefits accruing well into the future, the choice of discount rate had the greatest impact on results, particularly for the HPVV. There is considerable uncertainty in the underlying epidemiological and efficacy data, and estimates of incidence of disease, mortality rates, and vaccine efficacy were highly influential inputs affecting study findings. Vaccine program costs were highly influential parameters, particularly in Tuvalu, where the cost per DALY averted approached 2× GDP per capita for PCV and exceeded 3× GDP per capita for RVV at the higher parameter estimates. This finding is largely driven by the high estimated costs of immunisation programs in Tuvalu, where the population is smaller and more geographically dispersed on outer islands compared to the other three countries. Cost-effectiveness ratios for HPVV were favourable across most parameter estimates evaluated, falling just over, or well below the lower estimate of willingness to pay thresholds in each country. Payer perspective (government or societal) and vaccine coverage estimates had the least effect on cost-effectiveness for all vaccines in all countries.

**Figure 2:**
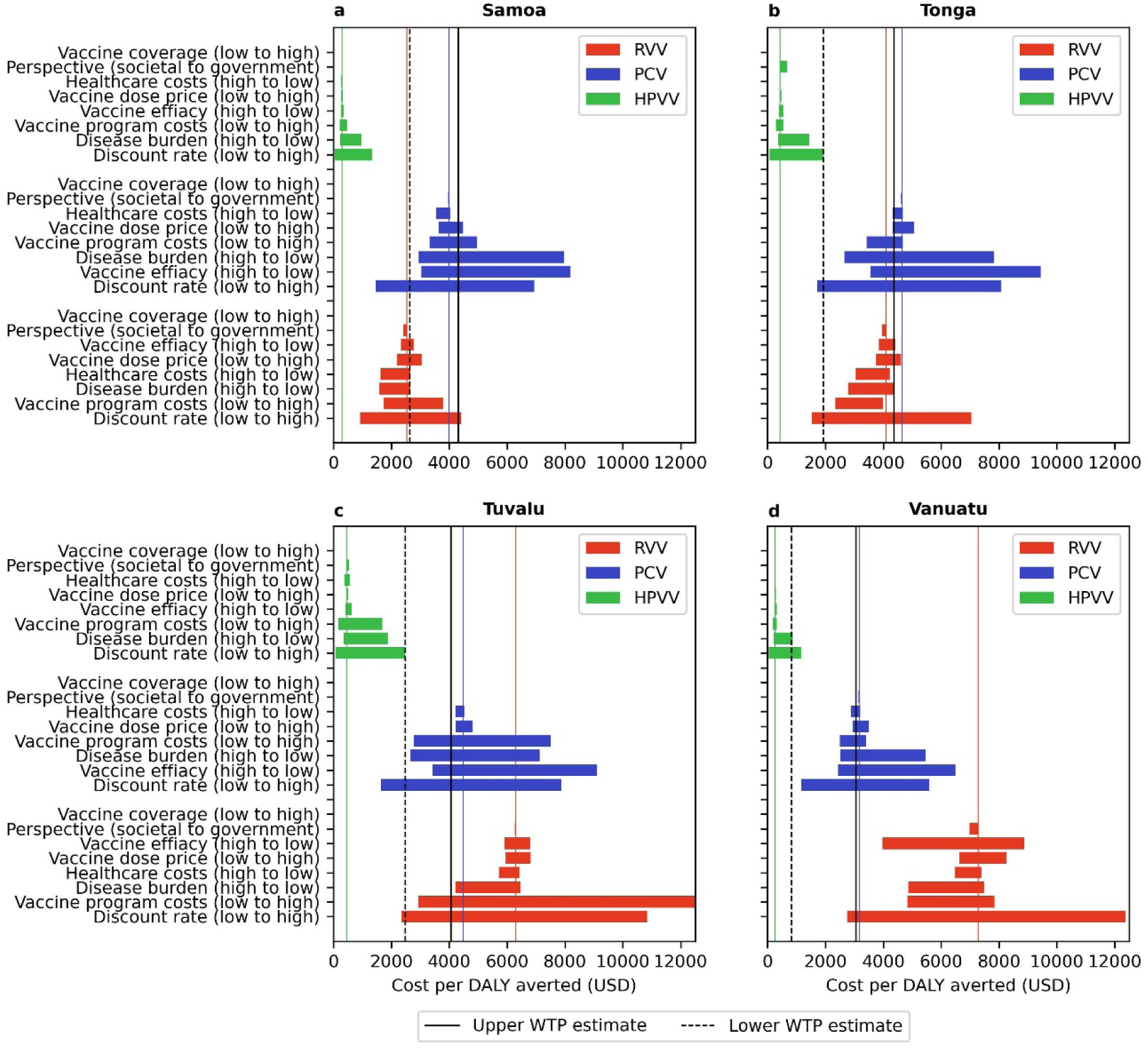
One-way sensitivity analysis of the cost effectiveness ratios for HPVV, PCV and RVV in a: Samoa; b: Tonga; c: Tuvalu; and d: Vanuatu. Disease event rates include upper and lower estimates for both incidence and mortality. Vaccine efficacy includes upper and lower estimates for both efficacy and duration of effect. Solid coloured lines represent base case cost-effectiveness ratio for each vaccine. Black vertical lines represent WTP estimates. Upper WTP estimate is 1× GDP per capita in each country. Lower WTP estimate is 0·61×, 0·44×, 0·61×, and 0·21× GDP per capital in Samoa, Tonga, Tuvalu and Vanuatu, respectively. DALY: disability-adjusted life year; HPVV: Human papillomavirus vaccine; PCV: Pneumococcal conjugate vaccine; RVV: Rotavirus vaccine; USD: United States Dollars; WTP: Willingness to pay

The acceptability curves from the probabilistic sensitivity analyses are shown in Figure 3 and Webappendix D (Figure D3) for lower price vaccines. At the lower estimate of willingness to pay, HPVV is highly likely to be cost effective in all countries (probabilities >0·99), while the probability of PCV and RVV being cost-effective at this willingness to pay threshold is much lower (<0·006) in all countries except Samoa, where the probability of RVV being cost-effective is 0·877. At this willingness to pay threshold, simultaneous introduction of all three vaccines has a probability of being cost-effective ranging from 0·149 in Tuvalu to 0·973 in Samoa. If willingness to pay is 1× GDP per capita, the probability that introduction of all three vaccines is cost-effective is greater than 0·846 in all countries.

**Figure 3:**
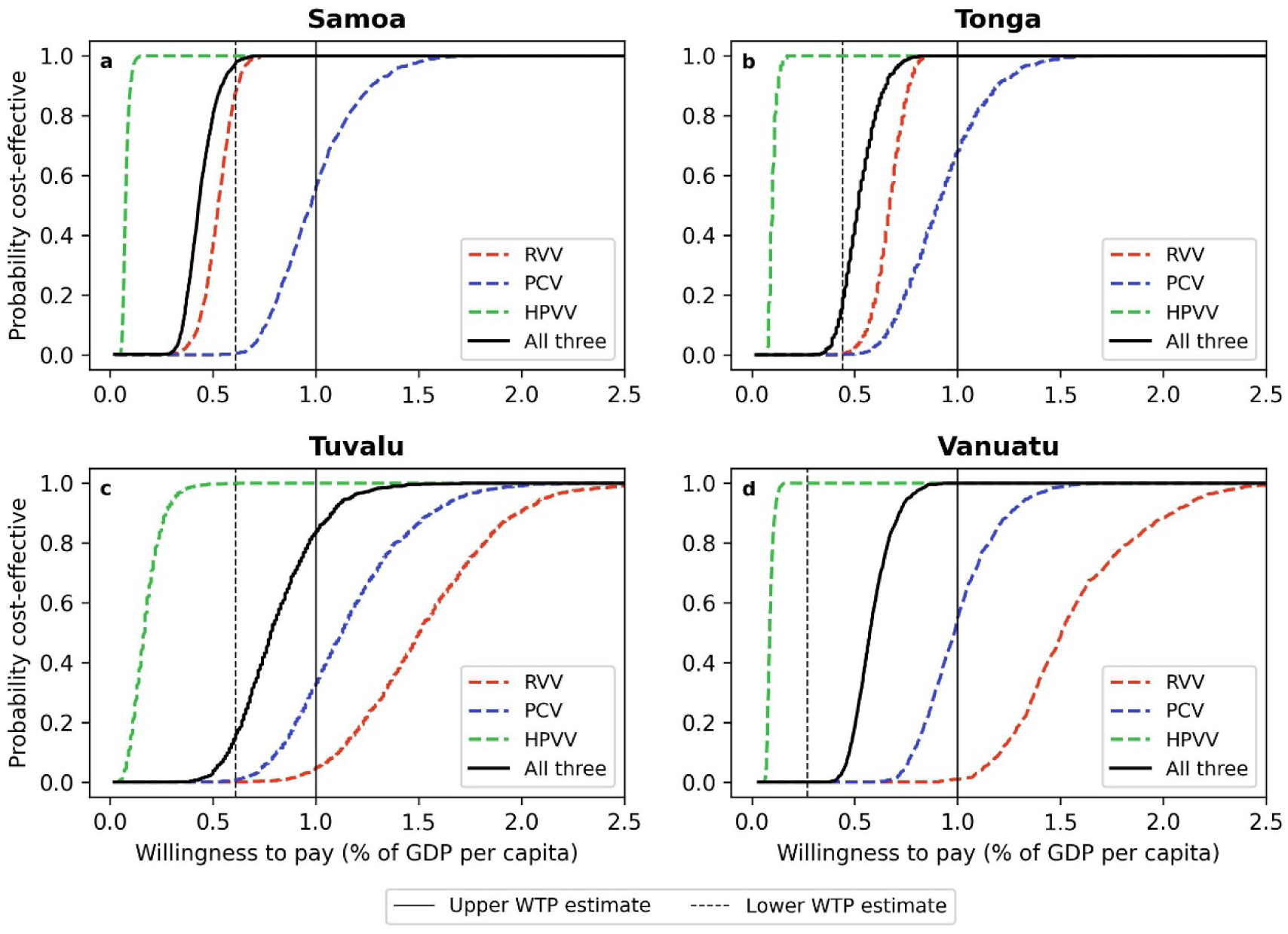
Cost-effectiveness acceptability curves for a: Samoa; b: Tonga; c: Tuvalu: and d: Vanuatu. Vertical lines represent WTP thresholds as a percentage of per capita GDP. Upper WTP estimate is 1× GDP per capita in each country. Lower WTP estimate is 0·61×, 0·44×, 0·61×, and 0·21× GDP per capital in Samoa, Tonga, Tuvalu and Vanuatu, respectively. GDP: Gross domestic product; HPVV: Human papillomavirus vaccine; PCV: Pneumococcal conjugate vaccine; RVV: Rotavirus vaccine; USD: United States Dollars

### Budget impact

Results from the budget impact analysis are shown in Figure 4 for the vaccines at PAHO Revolving Fund pricing and Webappendix D (Figure D4) for the lower price vaccines.

**Figure 4:**
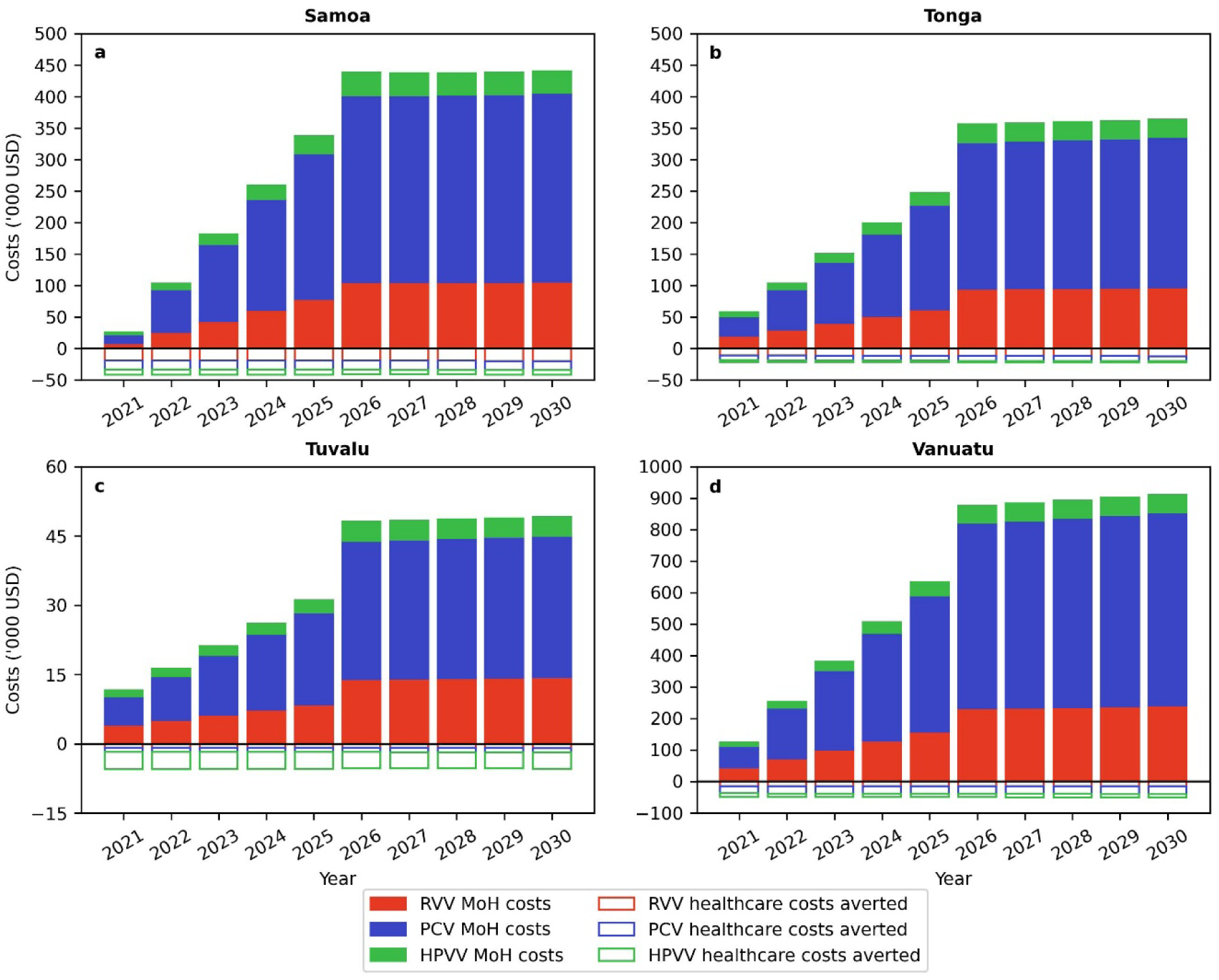
Results from budget impact analysis for HPVV, PCV, and RVV in a: Samoa; b: Tonga; c: Tuvalu; and d: Vanuatu. HPVV: Human papillomavirus vaccine; MoH = Ministry of Health; PCV: Pneumococcal conjugate vaccine; RVV: Rotavirus vaccine; USD: United States Dollars

Addition of the three vaccines would increase the cost of the immunisation program by 438,098 USD, 355,744 USD, 48,029 USD and 875,589 USD in Samoa, Tonga, Tuvalu and Vanuatu respectively after scaling up the full cost of the program to the government at year six. This would represent an increase in total health expenditure ranging from 0·45% in Tuvalu to 2·76% in Vanuatu each year. The annual increase in spending on vaccines would range from 359% in Samoa to 1368% in Vanuatu. When considering the reduction in spending on healthcare (by reducing disease burden), the increase spending on vaccines would range from approximately 326% in Samoa to 1290% in Vanuatu. If lower price vaccines were introduced, the annual impact on government budgets (at year six) would be 182,254 USD, 217,013 USD, 39,177 USD and 496,185 USD for the governments of Samoa, Tonga, Tuvalu and Vanuatu, respectively (representing an increase in vaccine budgets ranging from 149% in Samoa to 775% in Vanuatu).

## Discussion

This is the first study to provide evidence on the cost-effectiveness of three new vaccines under consideration for introduction into routine immunisation schedules simultaneously in four Pacific Island countries, with development partner support. The face validity of our findings can be demonstrated in their broad consistency with other analyses of the cost-effectiveness of these vaccines individually in these countries, in that the HPVV may be considered highly cost-effective,^66^ but the cost-effectiveness of RVV is highly dependent on price and unlikely to be favourable when the higher priced vaccine is considered individually.^67^ However, simultaneous introduction of all three vaccines was the policy decision these countries were faced with when this work was commissioned as the high financial and human cost of negotiating budgets, prices, and introduction activities in these settings made it necessary to consider introduction of all three vaccines or no vaccine at all. The findings of our study allow for consideration of the value of concurrent introduction of multiple vaccines, enabling favourable pricing negotiations and efficiencies in introduction activities. Our analysis found that introduction of HPVV, RVV and PCV together at PAHO Revolving Fund prices has a cost-effectiveness ratio of around half to three quarters of GDP per capita in each country, suggesting these vaccines represent good value for money based on thresholds historically recommended by the WHO Commission on Macroeconomics and Health for assessing cost-effectiveness based on a human capital approach (1× GDP per capita per DALY averted).^68^ This evidence formed part of a package of information guiding country-level decisions to introduce all three vaccines in these four countries. This study reflects the limited opportunity for introduction of all three vaccines together both due to logistics, operational support and the available funding window, which differs from, for example, Gavi-eligible countries. While the initial costs of introduction (purchase of vaccine doses and community mobilisation costs) were offset by financing support from ADB and UNICEF, all four country governments committed to progressively transitioning to 100% financing of the vaccination program.

Recognising new empirical global evidence suggesting that 1× GDP per capita may be too high a willingness to pay threshold, particularly for low- and middle-income countries, we also compared cost-effectiveness ratios to more recent estimates of willingness to pay that consider the opportunity costs of investing in a health intervention (and the benefits forgone by not investing those resources elsewhere).^62^ Based on these thresholds (available for Samoa, Tonga and Vanuatu), introduction of these three vaccines is still likely to be cost-effective in Samoa, but may exceed willingness to pay in Tonga, Vanuatu, and likely Tuvalu (although a willingness to pay threshold in the latter has not been reported). The lower price vaccines have a much higher likelihood of being cost-effective even at these more conservative estimates of willingness to pay, with costs per DALY averted ranging from 0·05× GDP per capita in Samoa to 0·58× GDP per capita in Tuvalu when all three vaccines are introduced. However, it should be noted that some lower-priced vaccines place an increased burden on immunisation programs (e.g. the lower priced RVV requires a three-dose schedule and is presented in a multidose vial, leading to higher wastage and requiring more resources for delivery).

Our analysis estimated that vaccine adoption would require substantial increases in vaccine budgets ranging from 349% in Samoa to over 1300% in Vanuatu. Even with the adoption of lower-priced products, vaccine budgets would need increases of between 149% (Samoa) to 775% (Vanuatu). These findings highlight that cost-effectiveness should be considered alongside affordability assessments and that fair pricing principles are paramount even if cost-effectiveness profiles are favourable. The introduction of vaccines has been slow in resource-limited countries which are not eligible for Gavi funding support, with vaccine price being a major barrier to introduction.^69^ Through pooled procurement, the Vaccine Independence Initiative has facilitated access to vaccine prices that are more affordable than what countries with small populations may be able to negotiate on their own terms. This has been reflected in our analysis, and higher dose prices with less favourable cost-effectiveness results may be expected if only a subset of vaccines were adopted by a few countries. However, more needs to be done to ensure middle-income countries, including small island countries in the Pacific, have access to affordable vaccines that don’t rely on partner assistance to introduce.^70^ The most recent PAHO Revolving Fund list (2024) includes recently pre-qualified vaccines from Indian manufacturers, with prices within the ranges used for our analysis of lower-cost vaccines. Enabling Pacific Island countries to access vaccines at these prices will be important to reduce the budget impact and strengthen the cost-effectiveness profile of PCV, RVV and HPV in these countries and others which are yet to adopt these vaccines.

There are limitations to our analyses that should be noted. There is considerable uncertainty in the underlying epidemiological and cost data available and used in this analysis. For many disease burden inputs we drew on data from Fiji, where surveillances systems are comparatively more developed, enabling access to a larger and more robust body of evidence than what is available for Samoa, Tonga, Tuvalu and Vanuatu. We consider this to be reasonable, as Fiji shares several health, demographic and economic features in common with the four countries of this analysis (such as similar GDP per capita, population structure, and under five mortality rates). Similarities in results between countries will, in part, be driven by reliance on common data for modelling disease burden. Likewise, differences can be attributed to a smaller number of inputs where country-specific data were used (such as case fatality ratio for severe pneumonia and diarrhoea). Use of country-specific data for these inputs was particularly important as geographical challenges with accessing timely health care may vary across these four countries which is reflected in the variation in under five mortality rates ranging from 11 per 1,000 live births in Tonga to 23 per 1,000 live births in Vanuatu. Disease incidence and mortality estimates were among the most influential parameters impacting cost-effectiveness ratios. Improved epidemiological data would provide more certainty on vaccine cost-effectiveness and improve capacity for evidence-based decision making in these countries. Countries can use the opportunity of development partner support to improve routine data collection and reporting and strengthen laboratory capacity.

We make several assumptions and model design choices that result in conservative estimates of the cost-effectiveness of these vaccines. The discount rate used in our base case analysis is conservative (3% for both costs and benefits) compared to current WHO recommendations of leaving benefits undiscounted while costs are discounted at 3% (which we used as the lower bound for our sensitivity analysis).^63^ The discount rate chosen for the base case was between the current WHO recommended rate and the funder’s recommended rate (6% for both costs and benefits). Secondly, our use of WHO-CHOICE estimates for unit costs of hospitalisation and outpatient visits represents a conservative assumption, as these estimates often underestimate the true costs of these services.^71^ For example, a systematic review of the costs of management of pneumonia in young children suggests that outpatient visit costs for non-severe and severe pneumonia are approximately 66 and 51 USD (2013) respectively,^72^ compared to WHO-CHOICE-based estimates of 7-8 and 14-17 USD (2019) respectively used in this study.^72^ Estimates of PCV and RVV coverage were based on current coverage of DTP-containing vaccines, which are well established in the immunization program of these countries with high coverage.^73^ Additionally, a relatively narrow range of coverage estimates were explored in the sensitivity analysis. Although this may overestimate the coverage of newly introduced vaccines, PCV and RVV would be given to infants at the same immunisation visit as DTP-containing vaccines, so any operational factors affecting delivery would be common across these vaccines. Further, recent data suggests that, a year after introduction, coverage of PCV and RVV was similar to that of DTP in all four countries.^73^

Modelling a three-dose schedule of PCV may overestimate the potential cost-effectiveness and budget impact in light of emerging evidence suggesting that a two-dose PCV schedule may offer comparable protection to a three-dose schedule. ^74,75^ It has been estimated that approximately 1.5 billion USD could be saved by optimising the PCV dosing schedule in the poorest countries.^76^ Finally, we do not consider indirect protection to unvaccinated people, resulting in a conservative estimate of the health impact and cost-effectiveness of vaccine introduction. PCV provides substantial indirect protection at high coverage as has been demonstrated in Fiji and elsewhere,^77–79^ which has been estimated to reduce incremental cost effectiveness ratios by 30-50%. Other studies have demonstrated other indirect effects, such as reductions in malnutrition following introduction of RVV.^14^ We acknowledge this evaluation captures a narrow set of benefits of these vaccines relative to the recently articulated Full Value of Vaccines Assessments framework, which calls for evaluation of broader social and economic benefits arising from vaccination programs beyond direct health outcomes.^80^

It should be noted that parameter estimates used in the model have been updated since 2019, when the results of this analysis were used as part of decision-making on the adoption of these vaccines. Updates were made to incorporate more robust data that have more recently become available. Updates included modelling a single dose schedule for both HPVV products considering recent evidence showing comparable efficacy between one and two doses. WHO has recommended one or two doses for both HPVV products modelled in this analysis (recommendations most recently updated in 2024 to include single dosing). Additionally, the health system costs of vaccine delivery were updated with recent country-specific estimates that are higher than those originally used but considered to more accurately reflect the high costs of these activities.^57^ Overall, these updates have resulted in more conservative estimates of cost-effectiveness.

Further limitations of our analysis should be noted. Firstly, the costs and health impacts of catch-up campaigns were not included in this evaluation. Vaccine-specific catch-up programs for certain age groups were under consideration for some of the countries alongside routine introduction but were ultimately not implemented. Next, while we consider the interactions of introducing multiple vaccines simultaneously in regards to efficiencies in healthcare infrastructure requirements and vaccine program costs, we do not consider any differential health impacts compared a single vaccine, as these are likely to be negligible. Further, we valued lost days of work based on formal labour force participation, which does not account for unpaid time and underestimates opportunity costs attributable to those not in the formal labour force but who are engaged in other forms of productive labour (including unpaid household labour). Finally, the limitations of using a static model should be acknowledged. Dynamic transmission models allow for capture of potential indirect effects, including both herd immunity and serotype replacement, which can be particularly significant for some vaccines. The recent literature indicates little difference between static and dynamic models in some circumstances, including when evaluating the cost-effectiveness of RVV with a focus on outcomes in children under 5 years, and HPV vaccination of adolescent girls. For PCV, the potential for indirect effects (herd immunity and serotype replacement) is much greater than for rotavirus and HPV vaccines, thus a dynamic model could demonstrate different results compared to a static model. While a recently published pseudo-dynamic model of PCV impact generated similar estimates to UNIVAC when using the same coverage scenarios, the study noted reduced potential for herd effects in LMICs compared to high income countries based on post-licensure data from a very small number of countries (Kenya, Gambia, Israel, England, Lao PDR, Mongolia), and the potential for serotype replacement, potentially making results more conservative.^81^ However, the LMICs with post-licensure data may not be representative of these Pacific Island nations. While we acknowledge that it remains unclear whether a dynamic modelling framework would make the results found in this study appear more or less cost-effective, we believe the findings are unlikely to vary significantly. Developing a dynamic transmission model and collecting the data necessary to calibrate it was beyond the scope of the commissioned analysis and would not have been feasible within the policy window when introduction of these vaccine was being considered (which occurred just prior to and during the COVID-19 pandemic). Our results demonstrate that, despite the existing uncertainty in some key model inputs, low-cost vaccines are likely to be cost-effective and the base case results with higher cost vaccines are potentially cost-effective, depending on willingness to pay.

## Conclusion

The findings from this study suggest that HPVV, RVV and PCV introduced together may be cost effective in Samoa, Tonga, Tuvalu and Vanuatu, depending on willingness to pay and vaccine pricing. This evidence helped inform decisions to adopt all three vaccines at prices close to PAHO Revolving Fund pricing in these four countries. Further cost savings would be possible with the adoption of lower priced vaccines that have recently become available, highlighting the importance of ensuring that these vaccines are accessible to middle-income countries ineligible for Gavi financing. However, this alone is insufficient justification to introduce new vaccines, which also need to be financially sustainable. Ensuring sufficient fiscal space for health programs to continue to meet the populations health needs and expectations requires Ministries of Health and of Finance to work closely together. However, it should be recognised that even with expanding immunisation budgets, the price of currently available vaccines may continue to be unaffordable for many small island developing countries without partner support to reduce financial barriers to initial introduction. This underscores the importance of considering affordability constraints alongside cost-effectiveness and the need for innovative pricing mechanisms to ensure ongoing sustainability.^82^

## Data Availability

All relevant data are within the manuscript and its Supporting Information files.

## Acknowledgements

We acknowledge valuable research assistance from Emma Veve, Deputy Director General, ADB Southeast Asia Department and Patrick Abraham, University of Melbourne. We also acknowledge the valuable advisory support of all our collaborators at the Vanuatu Ministry of Health (Acting Director General for Health, Dr. Sam Posikai Tapo) and the Tuvalu Ministry of Health, the Samoa Ministry of Health and the Tonga Ministry of Health. These stakeholders have provided valuable insights to guide selection of model inputs most appropriate for these countries where availability of country-specific data is limited.

## Authors’ contributors

IML and FR helped conceptualize the study and provided senior scientific support and oversight of the project. SA, TN, CW and MO contributed data, reviewed model inputs and provided an essential link between the study and policy questions. NC, AC, RR and KL were involved in the study design. NC, EW, VLO and AC were involved in the data modelling, study analysis and data interpretation. MJ, RR and KL helped interpret results and provided senior scientific support. NC wrote the first draft and all authors revised the manuscript and provided intellectual content. All authors had full access to all the data in the study and all authors had final responsibility for the decision to submit for publication. All authors approved of the final version of this article.

